# Living through the heat: How urban children and young people experience and envision healthier cities

**DOI:** 10.1101/2025.06.17.25329755

**Authors:** Constance Bwire, Rachel Juel, James Milner, Gabrielle Bonnet, Ana Bonell, Shunmay Yeung, Harshita Umesh, Roberto Picetti, Sudheer Kumar Kuppili, Jessica Gerard, Robert Hughes

## Abstract

Climate change is driving more frequent and intense heatwaves, posing growing risks to urban populations, particularly children and young people (CYP). This study examined how heatwaves affect the health, well-being, and daily lives of CYP across six rapidly urbanising cities: Accra, Kumasi, Ouagadougou, Port Harcourt, Manila, and Dar es Salaam. We conducted online surveys during both heatwave and non-heatwave periods, collecting 2,269 valid responses. Heatwaves were defined as periods when apparent temperatures exceeded the 90th percentile of historical monthly averages for at least three consecutive days, validated by national meteorological data. Non-heatwave periods were defined as days within the same month and city when apparent temperatures were less than or equal to the 10-year average daily mean. Participants, CYP aged 13-29 and parents of children under 18, were recruited via targeted digital advertisements on Meta and Google. Quantitative data were analysed using descriptive statistics, chi-square tests, and logistic regression to assess associations between heatwave exposure and self-reported health symptoms, well-being, and daily disruptions. Thematic analysis of open-ended responses identified community priorities for climate resilience. Heatwaves were associated with higher reports of headaches, low mood, anxiety or stress, not enough food, and increased reliance on family support. Adverse effects were more pronounced among younger and lower-income participants. Participants highlighted five priorities for climate-resilient cities: more green spaces, improved water and sanitation, cleaner environments, stronger health and education services, and greater youth participation in decision-making. The results highlight the growing burden of heat-related health symptoms and daily activity disruptions among CYP and youth-informed strategies to reduce the unequal impacts of extreme heat in urban areas.

## Introduction

The nexus of children and young people (CYP)’s health, cities, and climate change is critical for the sustainability of global populations and the planet, yet we know little about how they experience their changing urban environments, especially during extreme event (1). Children worldwide are significantly exposed to climate change related events, including heatwaves. A child born today risks living in a world up to four degrees Celsius warmer than the pre-industrial average, where they could face unprecedented exposure to heatwaves, with projected increases in childhood asthma and other health impacts due to deteriorating air quality (2), directly impacting their health and subjecting them to the cascading effects of climate breakdown that threaten food, energy, and water supplies.

At the same time, children benefit the most from reduction of greenhouse gas emissions, elimination of air pollution, and the reversal of global warming. *C*hildren represent over 30% of the 4 billion people living in cities today (3). By 2050, the number of urban dwellers is projected to reach 6.7 billion, approximately 70% of the world’s population (3). While cities are not solely responsible for global emissions, they play a pivotal role in mitigation due to their dense populations, high levels of consumption, and potential for implementing sustainable infrastructure (4).

CYP, who are in a critical stage of physical, cognitive, and emotional development, are particularly vulnerable to the impacts of climate change, including increased risks of anxiety, depression, and post-traumatic stress disorder following extreme weather events (2,5,6). In addition, climate-related disruptions to essential services such as nutrition and education can further compromise their development and have long-term consequences for their physical and mental wellbeing (7). Despite growing recognition of the disproportionate impacts of climate change on CYP, there remain few attempts to systematically and rigorously capture the impacts of environmental conditions on this vulnerable group in cities, in particular their experiences of extreme weather/environmental events like heatwaves. Our previous work (7) addressed this gap by conducting cross-city surveys to capture the lived experiences of CYP during heatwaves. Building on these findings, the current study aims to further expand and deepen this evidence base to support inclusive and responsive urban climate adaptation strategies.

This research employs a cross-sectional survey method to collect and analyze data from CYP in six cities across diverse geographic regions experiencing heatwave and non-heatwave periods. By focusing on their experiences during both events, this research seeks to illuminate the challenges heatwaves pose as well as possible opportunities for improving urban environments for CYPs. Our method involved two phases of data collection: during heatwave period events and after they have subsided, to compare and quantify the impacts of heatwave and non-heatwave periods. This study provides both quantitative and qualitative insights into how heatwaves disrupt daily life, impact health, and influence perceptions of urban resilience. The findings offer important implications for designing inclusive, youth-informed strategies in urban planning and climate adaptation.

## Methods

### Study Design and Setting

Building on our previous study (7), this research employed a cross-sectional design and an online survey to collect data from CYP living in cities during heatwave and non-heatwave periods. Recruitment was conducted online through targeted advertising on Google and Meta platforms, focusing on six cities: Ouagadougou (Burkina Faso), Port Harcourt (Nigeria), Kumasi (Ghana), Accra (Ghana), Manila (Philippines), and Dar es Salaam (Tanzania). These cities were chosen based on the presence of verified heatwaves during the research period, identified through automated environmental monitoring and confirmed by the research team.

The six cities were selected from a larger pool of 178 focal cities identified by the Children, Cities and Climate (CCC) Action Lab. The CCC Action Lab is a global research initiative led by the London School of Hygiene & Tropical Medicine (LSHTM) focusing on amplifying youth voices and informing equitable climate adaptation in rapidly urbanising settings. These 178 cities were chosen for their youthful populations, fast-paced urbanisation, and relevance to CCC partner networks (see S1 File). The six cities included here were among those where automated systems detected heatwaves during the study period, allowing for timely survey deployment and comparative analysis.

In each city, two survey rounds were conducted: the first during the heatwave, and the second once heatwave conditions had subsided (non-heatwave). This enabled direct comparisons of CYP experiences during heatwave and non-heatwave periods to assess differences in impact.

### Sampling and Participants

A three-stage sampling approach was employed to identify cities and to recruit participants during both heatwave and non-heatwave periods:

#### Stage 1: Heatwave Identification

To systematically identify heatwaves across the 178 study cities, we defined a heatwave as a period of at least three consecutive days during which both maximum and minimum daily apparent temperatures exceeded the 90th percentile of historical averages for that calendar month, based on data from the past ten years (2013–2023) (8,9). Real-time temperature data were retrieved via Open Meteo APIs (https://open-meteo.com/) and integrated with Airtable (https://www.airtable.com/), enabling continuous, automated monitoring across all cities. When cities met the heatwave threshold, Airtable generated alerts to flag potential heatwave and non-heatwave periods. These alerts were then manually reviewed and verified by the research team using publicly available meteorological sources, including national weather service reports.

#### Stage 2: Recruitment During Heatwave periods

Once a heatwave event was identified, participants in that city were recruited through paid advertisements (ads) on Meta and Google platforms. These ads were targeted toward two key groups: young people aged 13–29 years, and parents or guardians aged 18 years and older with children under 18. Eligibility criteria included residing in the city currently experiencing a verified heatwave event, having internet access, and the ability to complete an online survey. Participation was entirely voluntary and anonymous. Each recruitment campaign remained open until either 7 days or until 640 eligible responses were submitted per event, whichever came first (S1 Fig.).

#### Stage 3: Re-sampling During Non-Heatwave Period

The survey was repeated in each city following the end of the heatwave. In this study, “non-heatwave periods” conditions referred to the average daily mean apparent temperature for each city during the same month, based on data from the past 10 years (8,10). As with event identification, the re-sampling process was automated using Airtable, which accessed temperature data through the Open Meteo APIs. Monitoring for non-heatwave event conditions began after the completion of each event survey. Airtable continued to monitor temperatures daily until a suitable non-heatwave event period was identified. S1 Fig. shows a detailed workflow.

### Data Collection

Data were collected from 10^th^ September 2024 and 28^th^ January 2025 using a short, self-administered online survey hosted on Typeform (https://www.typeform.com/). The survey was promoted through paid advertisements on Meta and Google platforms (S2 File), targeting people in cities experiencing either a verified heatwave event or non-heatwave event conditions. Each survey ran for up to 7 days or until the required number of eligible responses calculated individually per city (S1 Table) was reached.

The survey took around five minutes to complete and included both multiple-choice and open-ended questions (S3 File). It asked participants about their general well-being (11,12), health symptoms, disruptions in daily routines, and how well they felt their city responded to extreme weather. An open-ended question invited suggestions for making cities healthier and more sustainable. No personal information was collected.

Although the tool was available in twelve languages (S3 File), only the official or most commonly spoken language of each participating city was used. Translations were first generated using ChatGpt generative AI, then reviewed and refined by native speakers. Each version was pre-tested and improved based on feedback from the research project’s Youth Advisory Group and early pilot participants. This helped ensure that the survey was clear, accessible, and easy to understand across different cultural and linguistic contexts (12,13).

The consent form was brief (see S3 File) and explained the purpose of the study, the type of data collected, and that participation was voluntary and could be stopped at any time. For children and young people aged 0-18 years, parents or guardians completed the survey on their behalf. For adults aged 18-29 years, individuals completed the survey themselves, providing their own consent. No personal or identifying information, such as names, was collected. All responses were anonymous and securely stored on password-protected servers accessible only to the research team. Participants who gave consent were directed to the survey. Those who did not consent were exited from the survey automatically.

### Data Analysis

Quantitative and qualitative data collected during heatwave and non-heatwave periods were analyzed to explore differences in health, well-being, daily activities, and participant perceptions. Quantitative analyses were conducted using IBM SPSS Statistics Version 30, while qualitative analyses were performed using NVivo Version 15.

#### Quantitative Analysis

Descriptive statistics were used to summarize demographic characteristics, health and well-being variables, physical activity length, and daily disruptions. Data were stratified by heatwave event and non-heatwave event conditions. For continuous variables such as age and child’s age, independent samples t-tests were used. Categorical indicators such as gender, reported health symptoms, and daily disruptions were analyzed using Pearson’s chi-square tests.

To assess the association between heatwaves and non-heatwave periods and health-related outcomes, binary logistic regression models were employed. Ordinal logistic regression was used for outcomes such as self-reported general health and sleep quality, which were measured on a five-point ordinal scale ranging from 1 (Very Bad) to 5 (Very Good). These models included event exposure (heatwave / non-heatwave), demographic covariates (age, gender, income, city), and heatwave conditions as predictors. Binary logistic regression was used to analyze dichotomous outcomes, including the presence or absence of health symptoms (e.g., headaches, anxiety, respiratory issues) and daily activity disruptions (e.g., missed work or school) (S6 & S9 Tables). These models controlled for key demographic and environmental covariates to evaluate the likelihood of adverse outcomes during heatwave compared to non-heatwave periods.

#### Qualitative Analysis

Open-ended responses on how cities could be improved were analyzed using a thematic analysis approach in NVivo Version 15. A five-step process guided the analysis: familiarization with the data through iterative reading and keyword searches; initial coding using both inductive and in vivo approaches; theme development by grouping related codes; review and validation using NVivo’s coding comparison tool to ensure consistency; and final theme definition supported by illustrative quotes. This approach allowed for the identification of common priorities shared amongst participants (S5 File for the detailed Data Analysis Protocol).

#### Data Management and Cleaning

All survey responses were securely stored on Typeform servers, which are hosted on Amazon Web Services infrastructure. Responses were automatically linked to Google Sheets in real time, allowing the research team to monitor data quality and participation during each heatwave / non-heatwave periods. Final datasets were downloaded and processed on encrypted, password-protected devices.

Confidentiality was maintained throughout the research. No personally identifiable information, IP addresses, or digital tracking data (e.g., cookies) were collected by the research team.

Survey data were checked for completeness, duplication, and formatting consistency. Translation to English was conducted where necessary. Duplicate responses were identified and removed using the "Identify Duplicate Cases" function in SPSS. Missing data were minimal due to the mandatory nature of most survey questions. Responses such as “I prefer not to say” or “I don’t know” were retained as valid and analyzed accordingly. Incomplete responses due to disqualification (e.g., age ineligibility) were not included in the final analysis.

## Results

Across the six cities studied, a total of 2,269 eligible and consenting participants completed the survey during heatwave and non-heatwave periods. Accra received the highest number of heatwave-related responses (486), followed by Manila (362) and Port Harcourt (312). Non-heatwave responses were highest in Dar es Salaam (228), while other cities recorded notably fewer responses (Fig. 1).

**Fig. 1:**
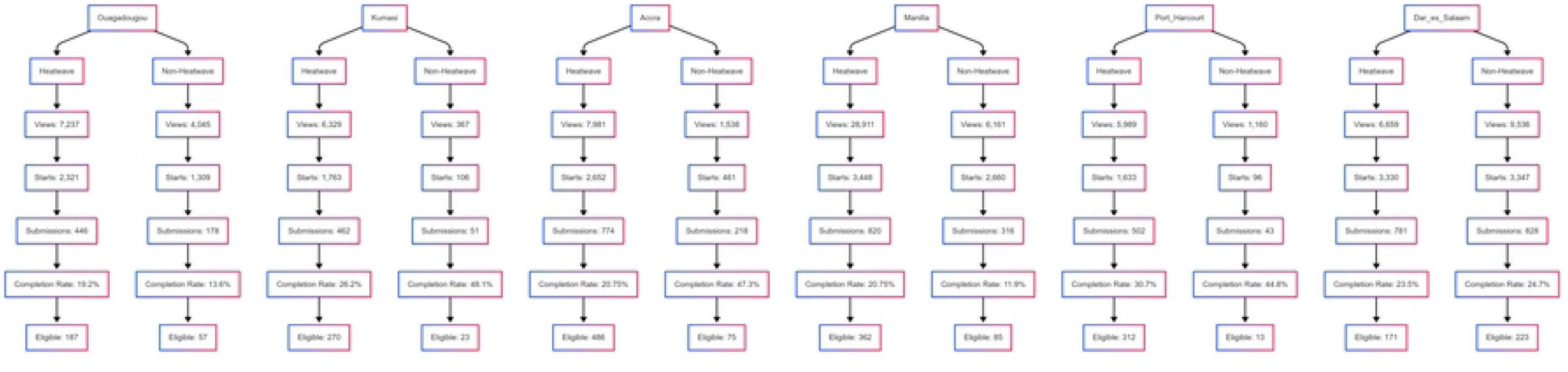
Survey participation by city and event status. Shows the number of eligible survey responses collected in each city during heatwave and non-heatwave periods.

Variation in participation volume and response quality was evident across cities and event conditions. Manila during the heatwave period recorded the highest survey engagement, with 28,911 views and 820 submissions, indicating strong public responsiveness during a heatwave. In contrast, Dar es Salaam during the non-heatwave period showed the greatest balance of scale and quality, with 828 submissions and 223 eligible responses. Kumasi during the non-heatwave period exhibited the most efficient response pattern, achieving a completion rate of 48.1% and a high proportion of eligible responses relative to total submissions (see Fig. 1 & S2 Table). Only a very small number of duplicates were identified, accounting for less than 0.5% of the total.

### Demographics Characteristics

Across all six cities (n=2269), just over half of the participants identified as parents (52.4%), with similar distributions observed during heatwave and non-heatwave periods. Gender representation was mostly males (57.8%), followed by females (39.7%), and a small portion (2.6%) who identified as other or preferred not to say. Most participants reported very low incomes (less than $100 / month) or were unsure of their income, highlighting widespread economic uncertainty.

The average age of participants was higher in the heatwave sample (31.5 years) compared to non-heatwave periods (28.9 years), with differences in average ages in Accra (younger participants during heatwaves) and Manila (older participants during heatwaves). The oldest participants overall were in Manila during heatwave periods (41 years). During heatwave conditions, the largest proportion of affected individuals falls within the 20-24 age group (30.8%), followed by 25-29 (23.3%) and 30-39 (16.6%). Conversely, younger individuals (1–14) and older adults (60+) show minimal representation at 0.2% and 0.6%, respectively.

In contrast, during non-heatwave conditions, the age distribution is slightly more balanced. The 20-24 group still represents the highest share at 31.0%, but the 25-29 group rises to 26.0%, with 30-39 decreasing to 14.8%. Notably, the older age group (60+) accounts for a slightly higher percentage than in heatwaves (1.5% vs. 0.6%) (See Fig. 2 for age distribution).

**Fig. 2:**
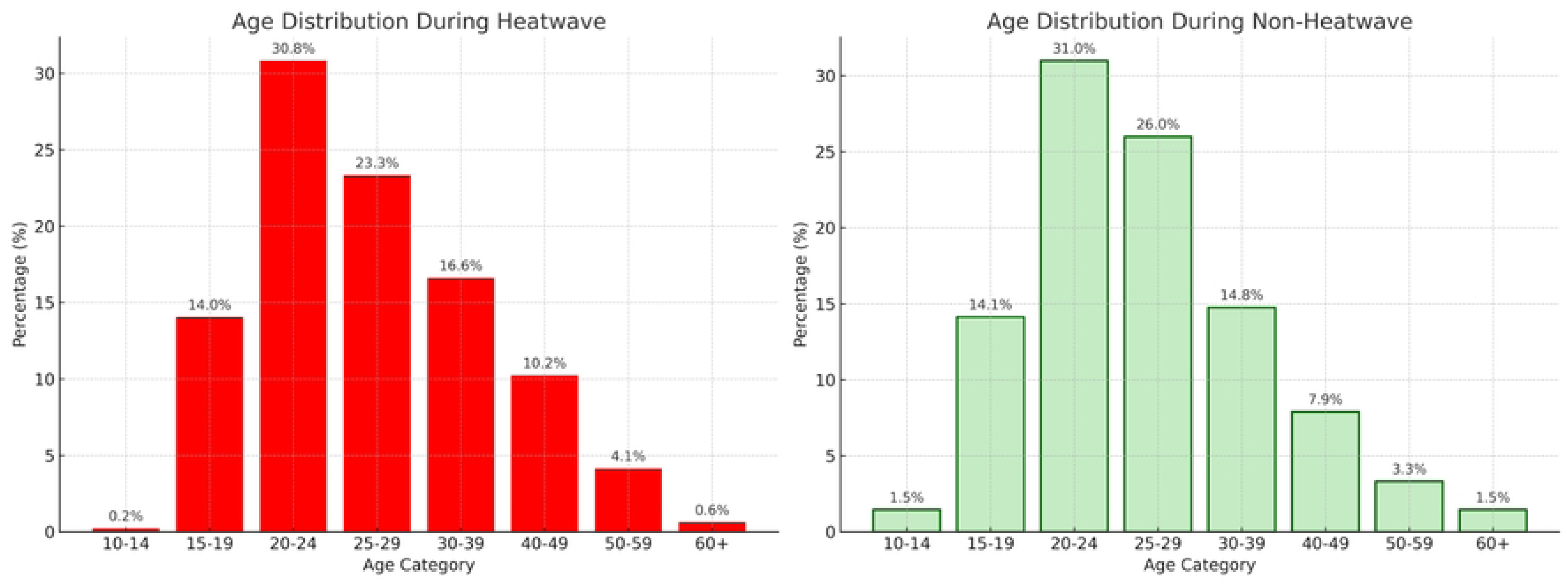
Age Distribution During Heatwave vs. Non-Heatwave Periods. Compares the age profiles of participants across both types of periods.

Among those who reported on behalf of a child, the average child age was reported as 6.9 years, and remained stable across conditions. Detailed demographic variables across the cities are provided in S3 Table.

### Perceived well-being and health symptoms

To assess general well-being, participants were asked about how they felt that day, how well they had slept the previous night, and any perceived health symptoms, both physical and mental.

Across all cities (n=2,269 total event sample; n=481 for heatwave periods and n=1,788 for non-heatwave periods) the proportion of participants feeling "Very good" was slightly higher during non-heatwave periods (53.6%) compared to heatwaves (50.7%), while responses of "Good" and "Ok" were similar between the two conditions. Reports of negative emotions ("Bad" or "Very bad") were more frequent during heatwaves (6.2%) than in non-heatwave periods (5.4%) (Fig. 3 & S4 Table). Overall, most participants reported positive general well-being under both heatwave and non-heatwave conditions.

**Fig. 3:**
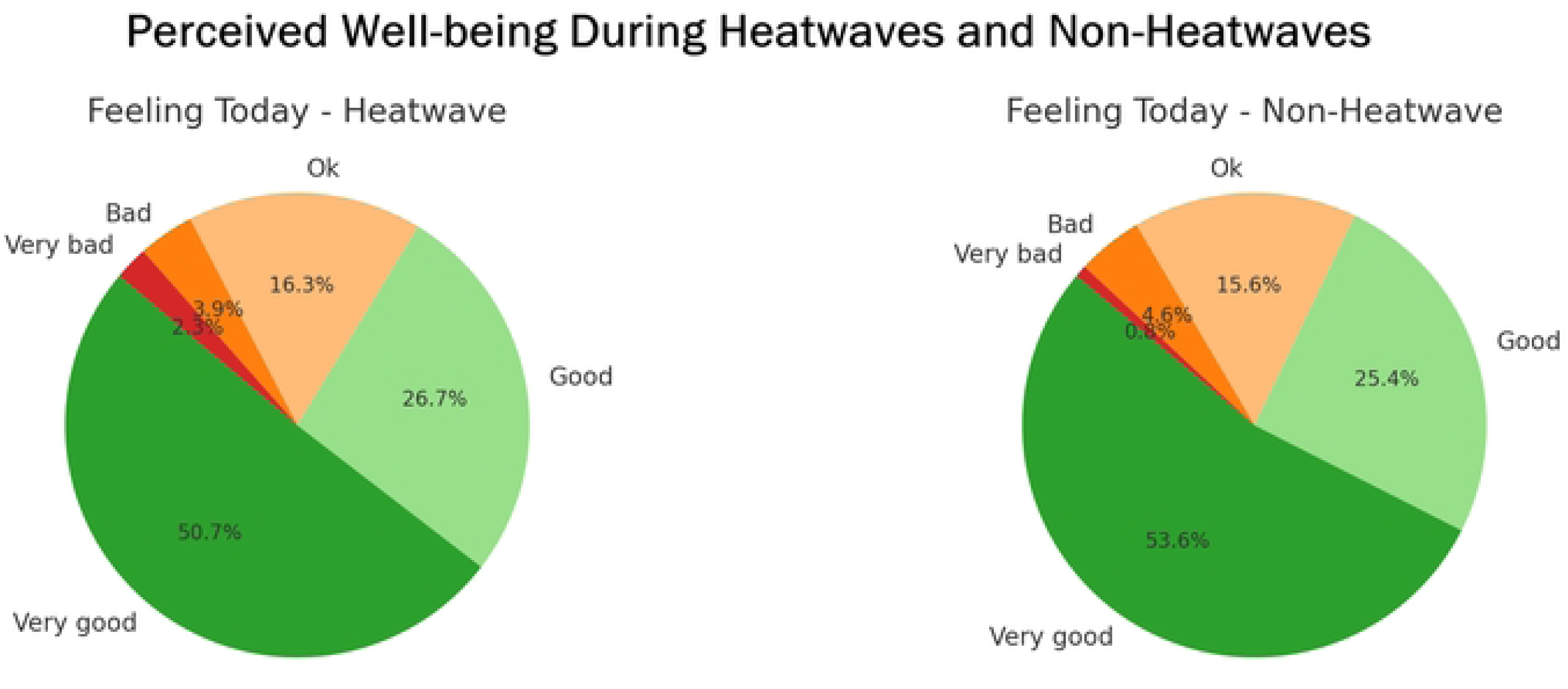
Perceived Well-being During Heatwave and non-heatwave periods. Presents how participants rated their general well-being at the time of the survey.

Across all cities, sleep quality followed a comparable pattern. The majority of participants reported "Very good" or "Good" sleep regardless of weather conditions, with only small shifts observed between groups. Reports of "Bad" sleep were slightly more common during non-heatwave periods (6.0%) than during heatwaves (5.1%), while "Very bad" sleep was reported more frequently during heatwaves (1.7%) compared to non-heatwave periods (0.8%) (Fig. 4 & S4 Table).

**Fig. 4:**
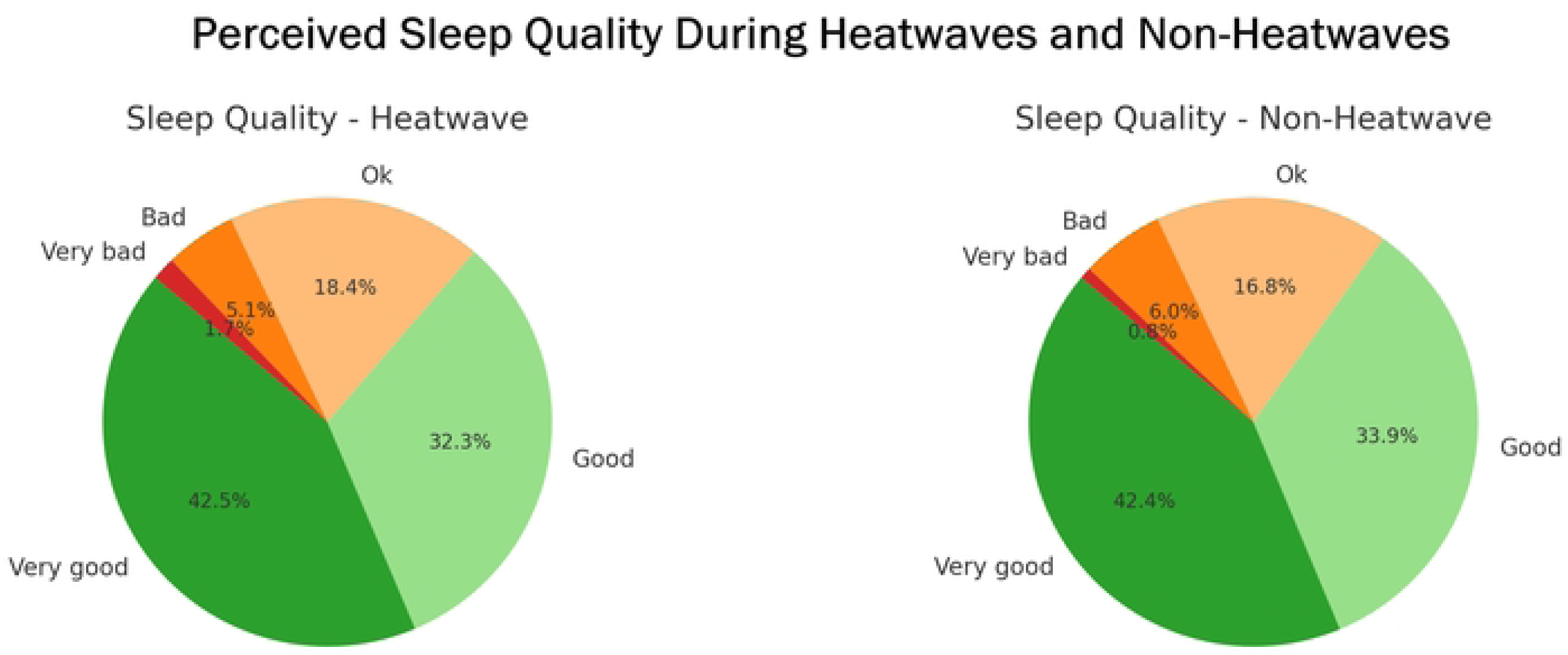
Perceived Sleep Quality During Heatwaves and Non-Heatwaves. Participants’ ratings of sleep quality from the previous night, shown by event type.

Participants across all cities reported higher levels of health symptoms during heatwaves compared to non-heatwave periods (S4 Table). Common symptoms such as headaches (44.1% vs. 37.8%), itchy eyes (30.1% vs. 27.2%), skin irritation (26.8% vs. 22.7%), and low mood (39.0% vs. 32.4%) were more frequently reported during heatwaves. Anxiety and stress were also more common (40.8% vs. 34.1%), while symptoms like diarrhea/vomiting and respiratory difficulties showed smaller differences between conditions (Fig. 5). City-level patterns varied: in Port Harcourt (n=325), heatwave-related symptoms were particularly high, with 53.5% reporting headaches, 33.3% skin irritation, and 45.2% low mood. Manila (n=447) also showed elevated symptom rates during heatwaves, including itchy eyes (27.9%) and anxiety/stress (41.6%). In contrast, Dar es Salaam (n=399) displayed more stable symptom reporting, with similar frequencies across both conditions. In contrast, the number of participants without symptoms was generally higher during non-heatwave periods (S5 Table).

**Fig. 5:**
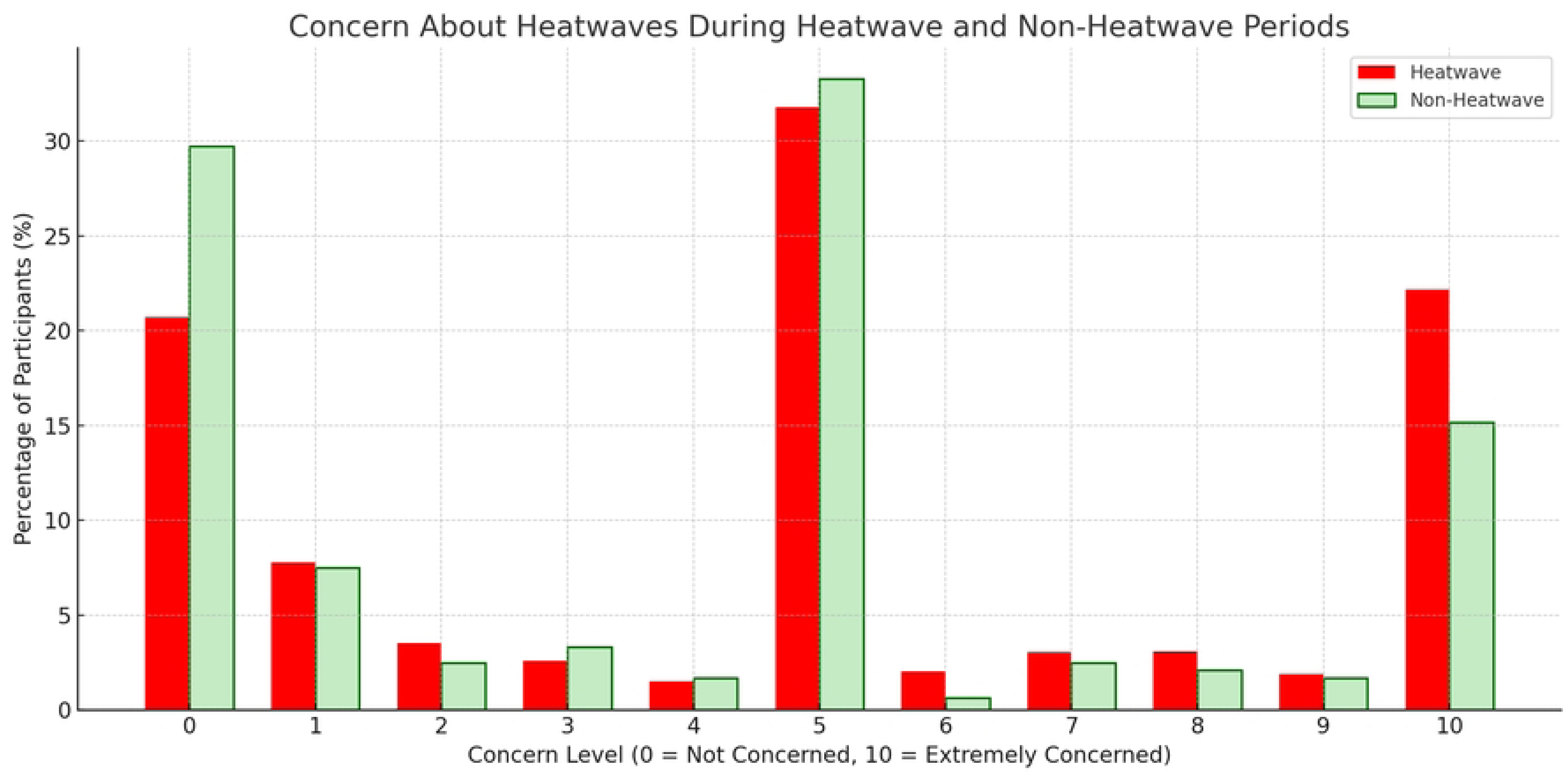
Perceived Health Symptoms During Heatwave and Non-Heatwave Periods. Participants reporting symptoms such as low mood, anxiety, and headaches during and after heatwaves.

Chi-square tests were used to examine the relationship between heatwave periods and reported well-being (feeling today and sleep quality) and health symptoms, both within individual cities and across all six cities combined. In Port Harcourt (n=325), heatwave periods were significantly associated with both how participants felt that day and their sleep quality (p < 0.001). In Dar es Salaam (n=399), only sleep quality showed a significant association (p = 0.002) (S4 Table). However, when data from all cities were pooled (n=2269), no significant link was found between heatwaves and well-being.

Health symptoms showed variation by city. In Manila (n=447), all reported symptoms were significantly associated with heatwave periods (p < 0.05). Ouagadougou (n=244) showed a significant result for heat exhaustion only, while Kumasi (n=293) showed no significant associations (S5 Table & S6 Table). In the pooled analysis, three symptoms, low mood (p = 0.008), anxiety or stress (p = 0.007), and headaches (p = 0.013), were statistically significant during heatwaves. These were selected for further analysis using logistic regression to examine their independent associations with heatwave exposure.

The regression models, across cities, showed that participants during heatwave periods were significantly more likely to report symptoms of low mood, anxiety or stress, and headaches (Table 1). Compared to participants in non-heatwave periods, those during a heatwave periods were 1.38 times more likely to report low mood (p = 0.003), 1.36 times more likely to report anxiety or stress (p = 0.004), and 1.33 times more likely to report headaches (p = 0.007) (Table 1 & S6 Table).

**Table 1:**
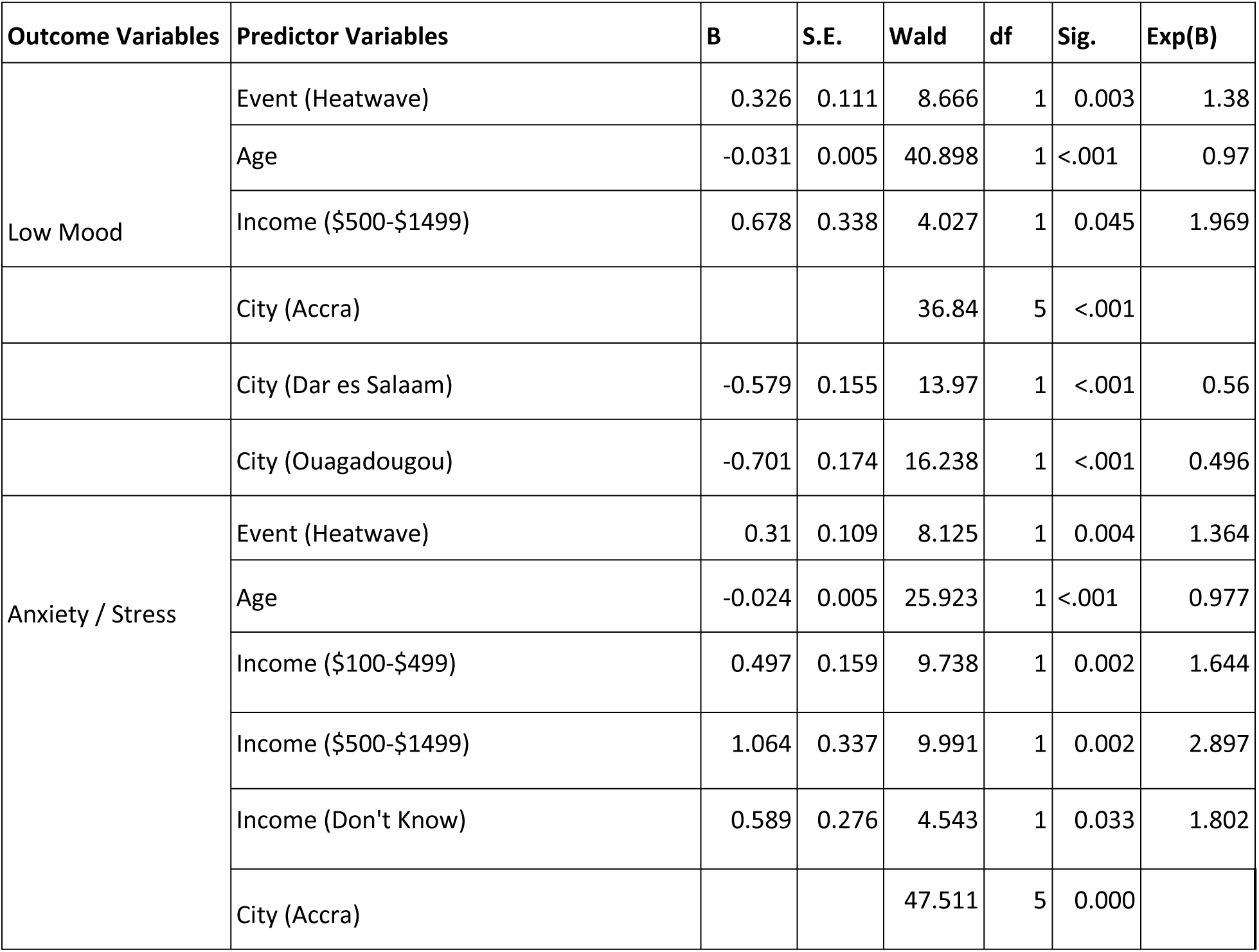
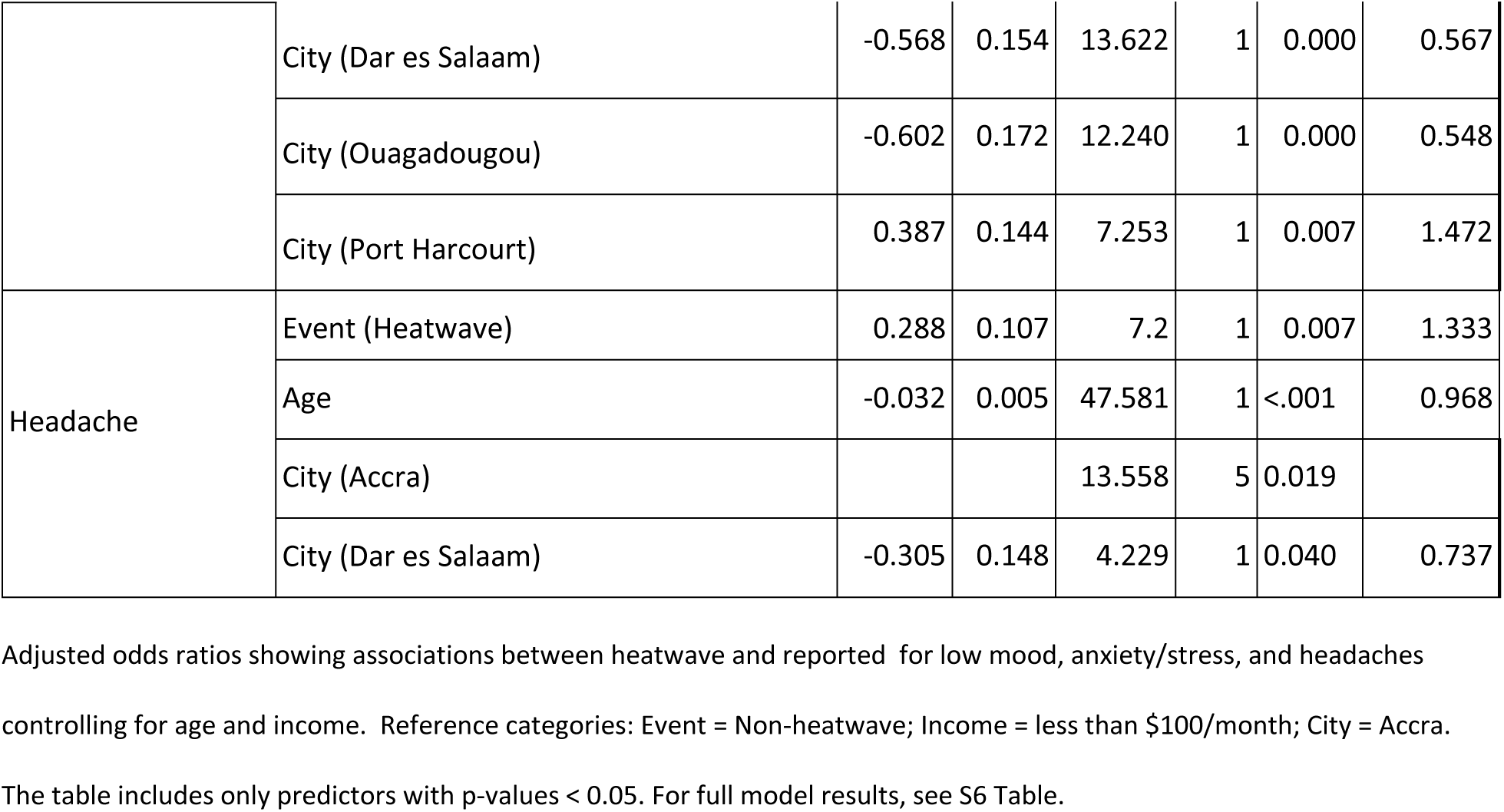
Logistic Regression Results for Health Symptoms During Heatwaves.

Participants’ younger age was significantly associated with higher odds of experiencing Low Mood, Anxiety or Stress, and Headache (OR = 0.97, *p* < 0.001). Regarding income, individuals earning $500–$1499 per month had increased odds of reporting Low Mood (OR = 1.97, *p* = 0.045) and Anxiety or Stress (OR = 2.90, *p* = 0.002), relative to those earning less than $100/month. Similarly, participants in the $100–$499 income group showed higher odds of Anxiety or Stress (OR = 1.64, *p* = 0.002) (Table 1).

Significant differences were also found between cities. Compared to Accra, participants in Dar es Salaam and Ouagadougou were significantly less likely to report low mood (OR = 0.56 and OR = 0.50) and anxiety/stress (OR = 0.57 and OR = 0.55). Participants in Port Harcourt were significantly more likely to report anxiety/stress (OR = 1.47, p = 0.007). For headaches, those in Dar es Salaam had significantly lower odds than those in Accra (OR = 0.74, p = 0.040). Although these city differences were statistically significant, the strength and direction of the associations varied.

### Disruptions to Daily Activities

Inactivity, i.e., 0 minutes of physical activity, was lower during heatwaves (14.5%) than during non-heatwave periods (16.8%). Meanwhile, a slightly higher proportion of participants reported engaging in more than 60 minutes of activity during heatwaves (20.4%) compared to non-heatwave days (19.8%). Port Harcourt (n=325) recorded high levels of extended activity in both periods, with 27.9% of participants engaging in more than an hour of physical activity during heatwaves and 30.8% during non-heatwave periods. Notably, no participants in Port Harcourt reported being completely inactive, i.e., zero minutes of physical activity in either event. In contrast, Dar es Salaam (n=399) had the highest rate of inactivity during non-heatwave periods (23.7%) and lower levels of extended activity, with only 11.7% reaching more than 60 minutes during heatwaves (S7 Table). Overall, physical activity levels were slightly higher during heatwaves than in non-heatwave periods.

Disruptions to daily activities were common across all cities, with some variables showing notable differences between heatwave and non-heatwave periods (S8 Table). For instance, Port Harcourt (n=325) experienced the most disruption during heatwaves across nearly all variables.

Notably, 57.9% reported not having enough food and 57.1% reported needing more family assistance (S8 Table). Overall, the proportion of participants reporting not having enough food was higher during heatwaves (44.1%) compared to non-heatwave periods (37.6%), as was the reported need for additional family assistance (46.3% vs. 40.1%). In contrast, some disruptions, such as being late for school or work (33.7% vs. 34.3%) and missing school or work completely (24.6% vs. 24.3%), remained relatively stable across conditions. Access to water and healthcare appointments also showed only minimal differences (Fig. 6).

**Fig. 6:**
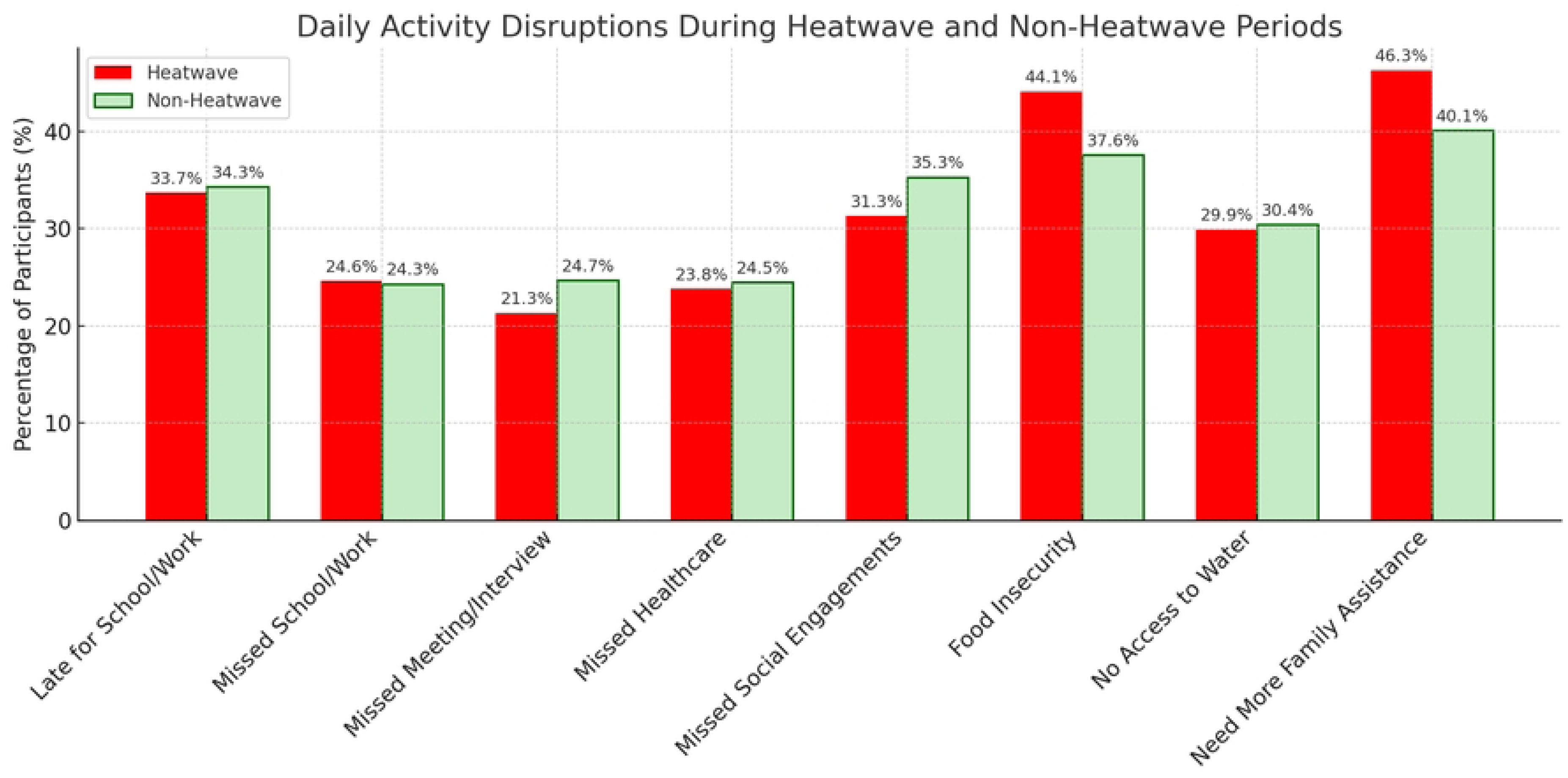
Daily Activity Disruptions During Heatwave and Non-Heatwave Periods. Reported daily disruptions such as missed work or school, and access to services.

Chi-square tests examined associations between heatwave periods and daily activity disruptions across cities. Several disruptions were significantly more common during heatwave periods. In Manila (n=447), all variables, including lateness, missed school or work, missed healthcare, missed interviews, and changes to daily routines, were significantly associated with heatwave periods (p < 0.05) (S8 Table). In Accra (n=561), significant associations were found for missed school or work, missed healthcare, lack of access to water, and increased need for family assistance. Dar es Salaam (n=399) showed a significant association for missed school or work only (S8 Table).

Across all cities, only two disruptions, not having enough food and needing additional family support, were consistently significant (p < 0.05) and were selected for further analysis using logistic regression (Table 2 & S9 Table).

**Table 2:**
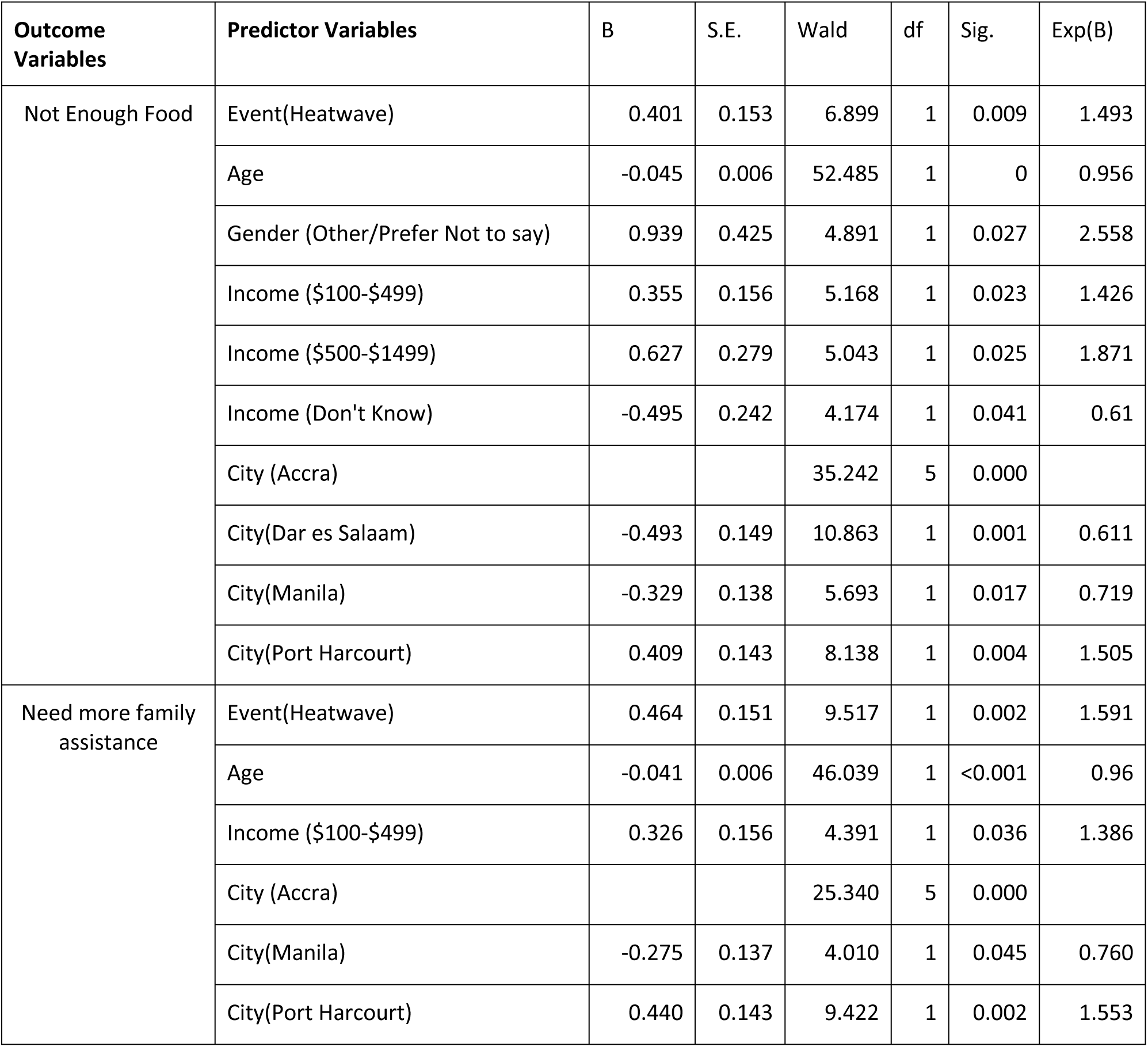
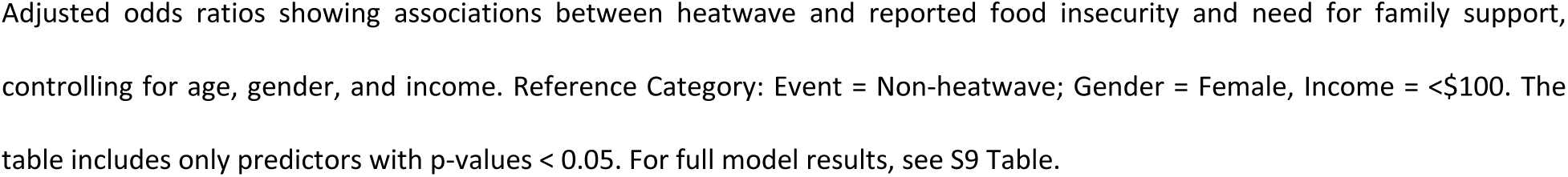
Logistic Regression Results for Disruptions in Daily Activities During Heatwaves.

| Outcome Variables | Predictor Variables | B | S.E. | Wald | df | Sig. | Exp(B) |
| --- | --- | --- | --- | --- | --- | --- | --- |
| Not Enough Food | Event(Heatwave) | 0.401 | 0.153 | 6.899 | 1 | 0.009 | 1.493 |
|  | Age | -0.045 | 0.006 | 52.485 | 1 | 0 | 0.956 |
|  | Gender (Other/Prefer Not to say) | 0.939 | 0.425 | 4.891 | 1 | 0.027 | 2.558 |
| | Income (\$100-\$499) | 0.355 | 0.156 | 5.168 | 1 | 0.023 | 1.426 |
| | Income (\$500-\$1499) | 0.627 | 0.279 | 5.043 | 1 | 0.025 | 1.871 |
|  | Income (Don't Know) | -0.495 | 0.242 | 4.174 | 1 | 0.041 | 0.61 |
|  | City (Accra) |  |  | 35.242 | 5 | 0.000 |  |
|  | City(Dar es Salaam) | -0.493 | 0.149 | 10.863 | 1 | 0.001 | 0.611 |
|  | City(Manila) | -0.329 | 0.138 | 5.693 | 1 | 0.017 | 0.719 |
|  | City(Port Harcourt) | 0.409 | 0.143 | 8.138 | 1 | 0.004 | 1.505 |
| Need more family assistance | Event(Heatwave) | 0.464 | 0.151 | 9.517 | 1 | 0.002 | 1.591 |
|  | Age | -0.041 | 0.006 | 46.039 | 1 | <0.001 | 0.96 |
| | Income (\$100-\$499) | 0.326 | 0.156 | 4.391 | 1 | 0.036 | 1.386 |
|  | City (Accra) |  |  | 25.340 | 5 | 0.000 |  |
|  | City(Manila) | -0.275 | 0.137 | 4.010 | 1 | 0.045 | 0.760 |
|  | City(Port Harcourt) | 0.440 | 0.143 | 9.422 | 1 | 0.002 | 1.553 |

Adjusted odds ratios showing associations between heatwave and reported food insecurity and need for family support, controlling for age, gender, and income. Reference Category: Event = Non-heatwave; Gender = Female, Income = <\$100. The table includes only predictors with p-values < 0.05. For full model results, see S9 Table.

The findings show that participants were 1.49 times more likely to report not having enough food (p = 0.009) and 1.59 times more likely to say they needed more assistance from family members (p = 0.002) when a heatwave occurred (Table 2).

Age was significantly associated with both outcomes. Younger participants were more likely to report not having enough food (OR = 0.956, p < 0.001) and a greater need for family support (OR = 0.96, p < 0.001). This group was 2.56 times more likely to report not enough food compared to women (p = 0.027). Income level showed statistically significant associations with disruptions to daily activities. Participants earning $100–$499 per month had significantly higher odds of experiencing Not Enough Food (OR = 1.43, p = 0.023) and Need More Family Assistance (OR = 1.39, p = 0.036). Those in the $500–$1499 income group also had significantly higher odds of Not Enough Food (OR = 1.87, p = 0.025) (Table 2).

City-level differences were significant for both outcomes. Compared to Accra, participants in Dar es Salaam and Manila were significantly less likely to report not having enough food during heatwaves (OR = 0.61 and OR = 0.72, respectively). In contrast, those in Port Harcourt were significantly more likely to report not having enough food (OR = 1.51, p = 0.004). For the outcome of needing more family assistance, participants in Manila were also less likely to report this need (OR = 0.76, p = 0.045), while those in Port Harcourt were significantly more likely to report needing family support during heatwaves (OR = 1.55, p = 0.002). (S9 Table for detailed analysis).

### Heatwave Concerns and Response Satisfaction

Across all cities, the most frequently selected concern levels during heatwaves were 5 (31.8%) and 10 (22.1%), representing over half of all responses (Fig. 7). In contrast, only 33.3% of participants chose these levels during non-heatwave periods. Nearly 30% reported no concern (0/10) when not experiencing a heatwave, highlighting a notable increase in concern with direct exposure. City-level patterns varied, but some, such as Accra and Manila, showed particularly high concern, with over 50% of participants selecting scores of 5 or higher. Others, like Dar es Salaam, had a larger share of respondents reporting no concern (53.2%) during non-heatwave conditions (S10 Table). Overall, most participants reported moderate to high levels of concern about heatwaves, particularly during periods of extreme heat.

**Fig. 7:**
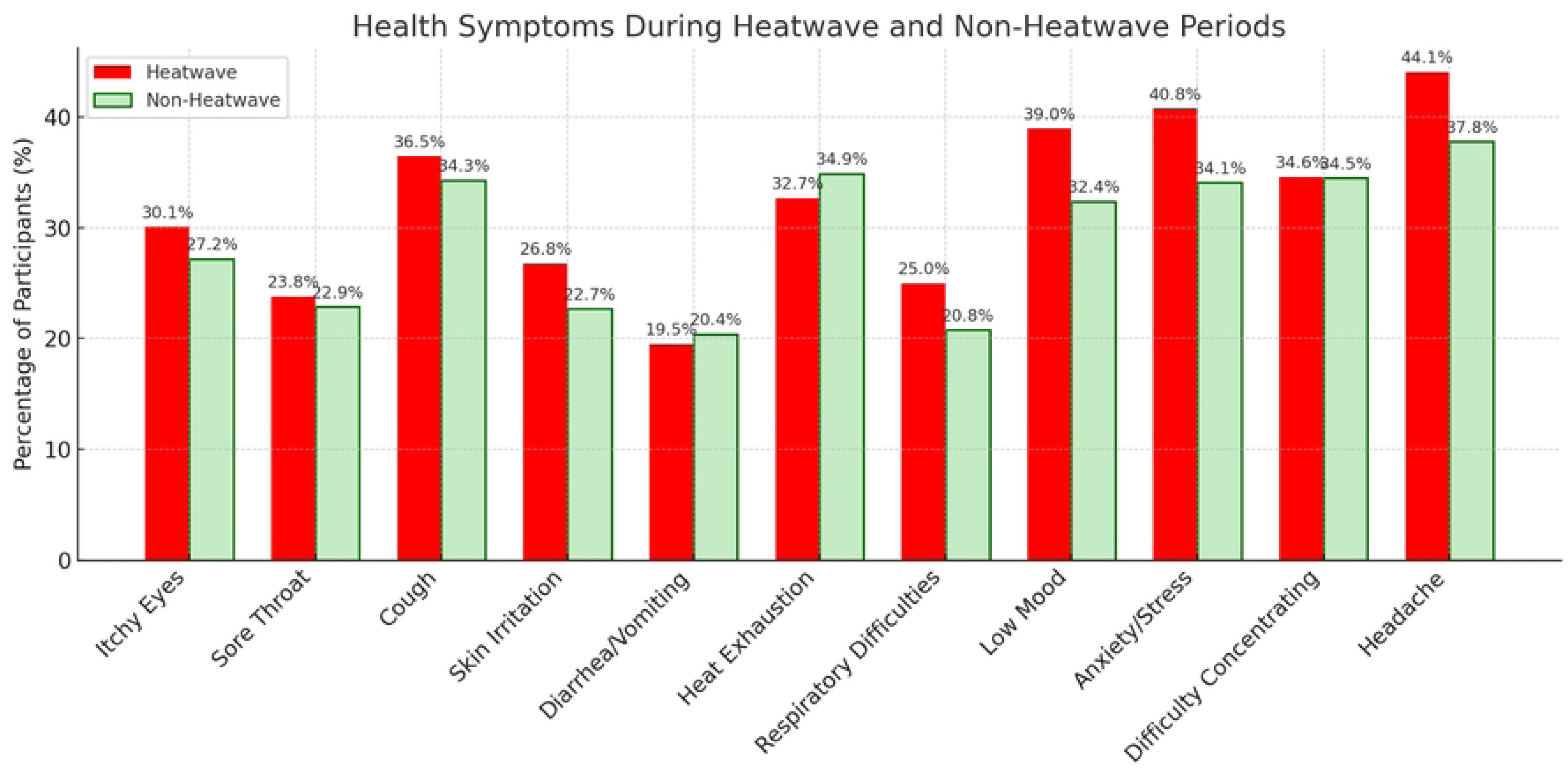
Concern About Heatwaves During Heatwave and non-heatwave periods. Participants’ concern about heatwaves, rated on a scale from 0 (no concern) to 10 (very concerned), by event type.

Participants across all cities reported a range of satisfaction levels with heatwave responses. During heatwaves, the most common response was “Very Satisfied” (34.6%), followed by “Neutral” (33.3%), while 19.5% expressed dissatisfaction. In non-heatwave periods, satisfaction was even higher, with 48.6% of participants reporting being “Very Satisfied,” and fewer choosing “Neutral” (26.0%) (Fig. 8 & S11 Table). City-level patterns varied. In Port Harcourt (n=325), 34.0% of participants were “Very Satisfied” during heatwaves, and only 11.0% reported any dissatisfaction. Dar es Salaam (n=399) showed even stronger approval, with 72.0% selecting “Very Satisfied.” Accra (n=561), on the other hand, had a more even spread of responses, including 34.0% “Neutral,” 26.3% “Very Satisfied,” and 25.3% reporting dissatisfaction (S11 Table

**Fig. 8:**
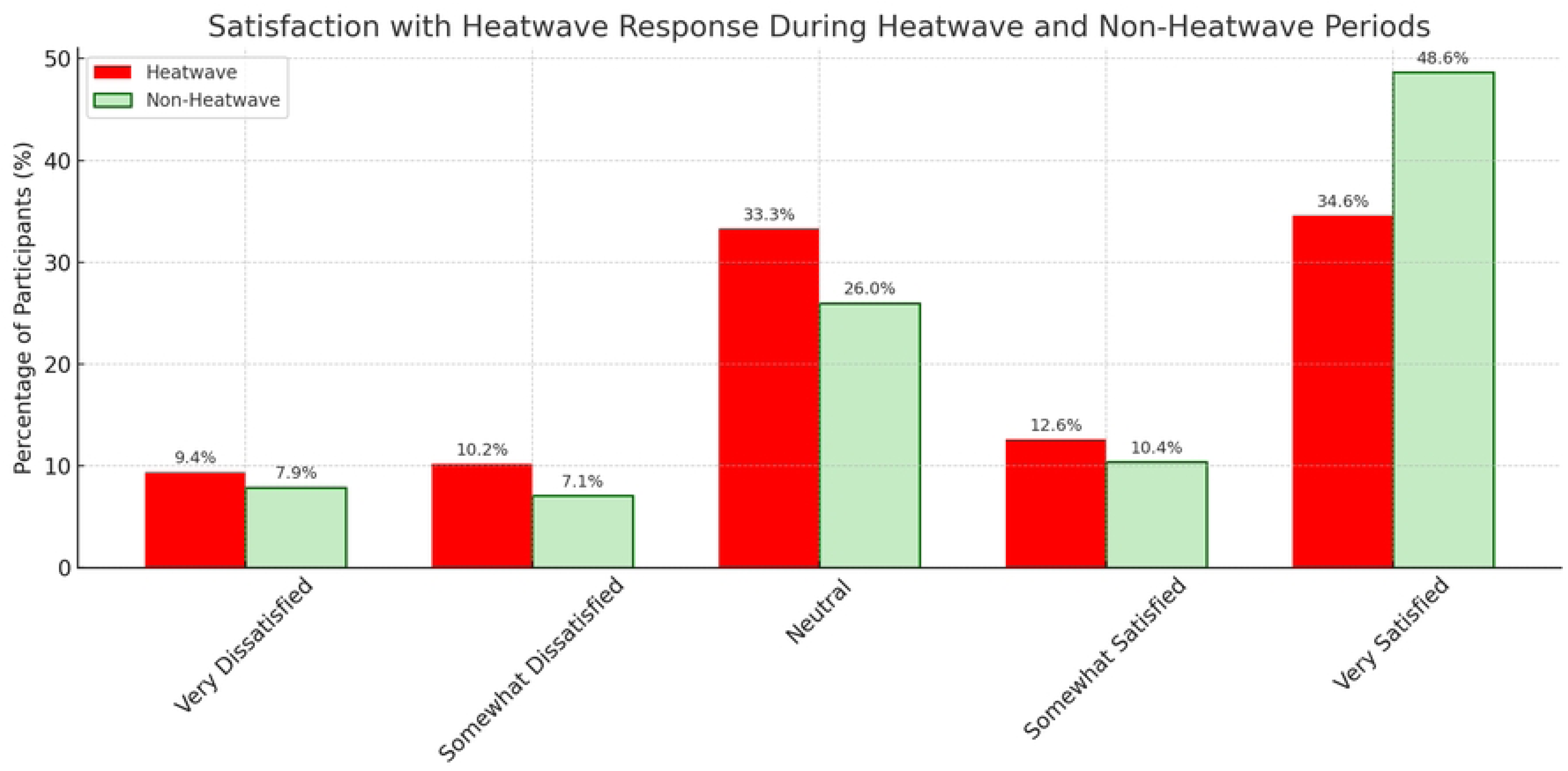
Satisfaction with Heatwave Response During Heatwave and Non-Heatwave Period. Participants’ views on how well their city responded to recent heatwaves, based on satisfaction level.

### Suggestions for Healthier & Sustainable Cities

In response to the open-ended question, *’If you could do one thing to make your town/city healthier and more sustainable during heatwaves, what would it be?’*, the most frequent keywords from participants’ emphasized priorities around environmental sustainability and public health. Common words included “clean”, “trees,” “plant,”” “recycling,” “water,” “sanitation,” and “health.” These reflect strong community interest in greener environments and improved basic services (Fig. 9).

**Fig. 9:**
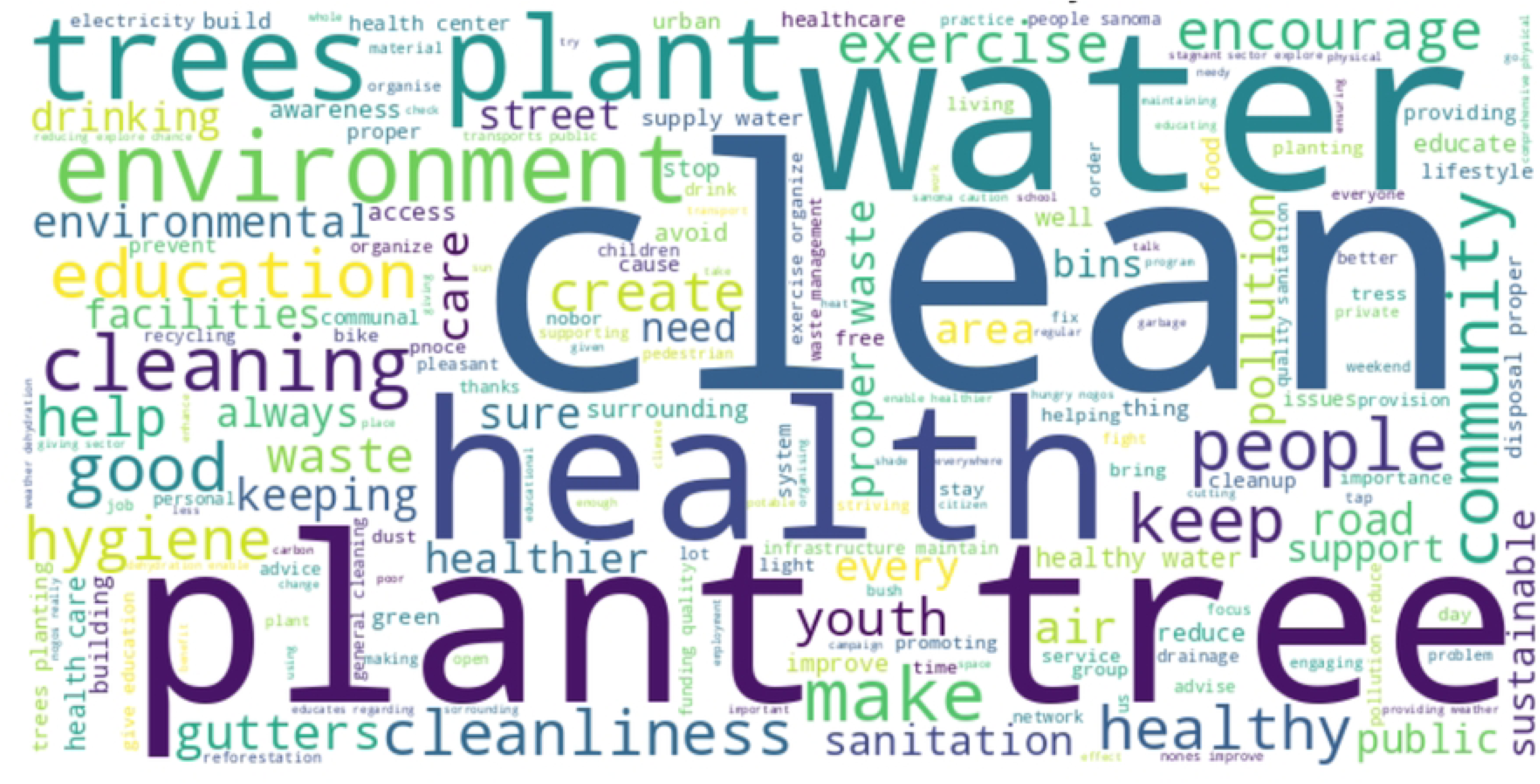
Word cloud of the most frequent keywords from participants’ suggestions for healthier and sustainable cities. Word cloud showing the most frequent suggestions from participants on how to improve their cities during heatwaves.

A thematic analysis of participant responses identified five key priorities for healthier urban living: Urban Greening, Water and Sanitation, Clean and Healthy Environment, Health Education and Services, and Civic Engagement. These themes, illustrated in Fig. 10 and expanded in (S12 Table), reflect actionable ideas rooted in community experiences and aspirations.

**Fig. 10:**
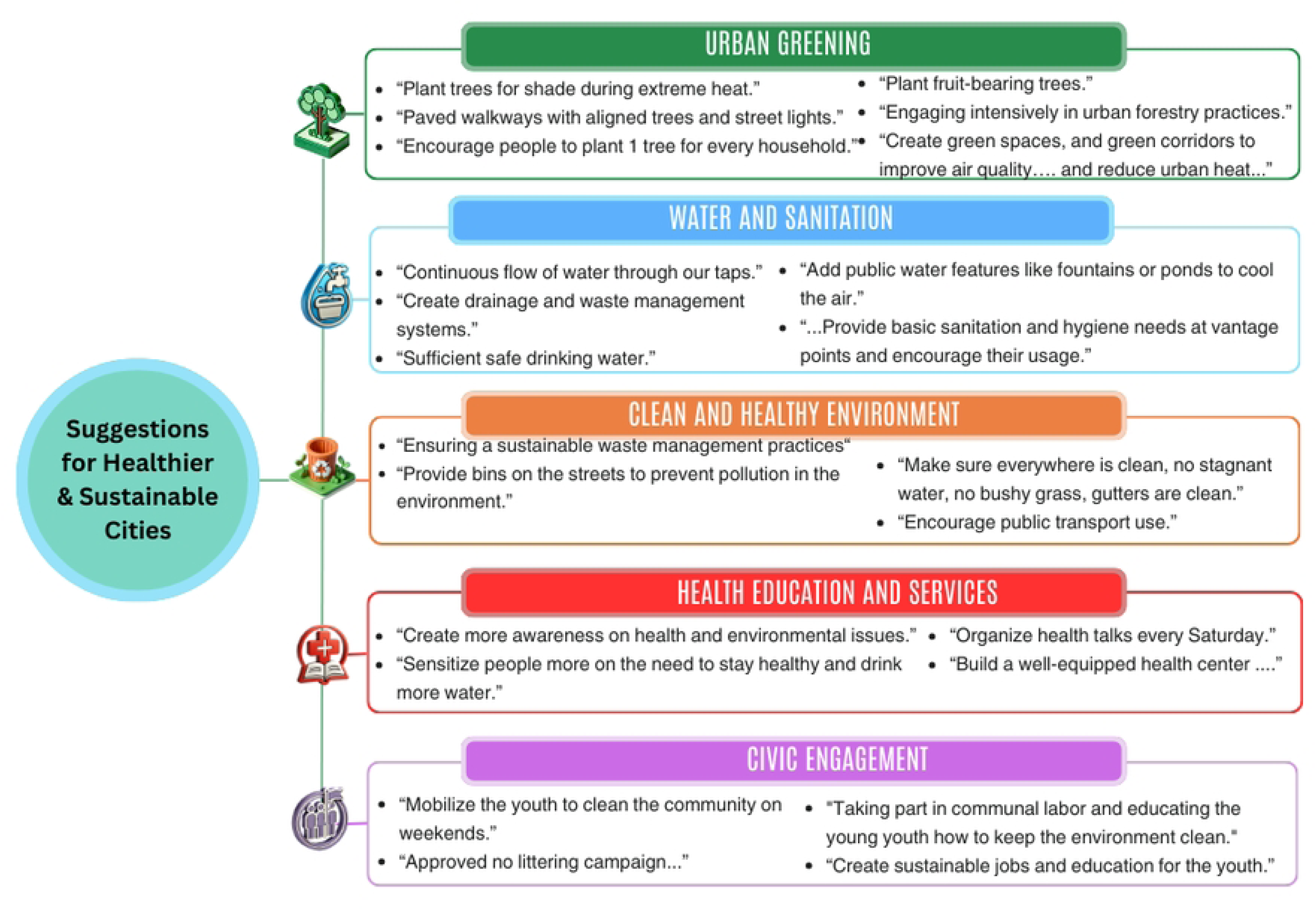
Themes, and illustrative quotes from participant responses on making cities healthier and more sustainable. Key themes from participants’ open-ended responses i.e., urban greening, water and sanitation, clean environments, health education and services, and civic engagement, along with selected example quotes from youth and caregivers.

Participants’ responses to the open-ended question revealed five main themes. Urban greening was the most common, with suggestions to expand green spaces and improve air quality, such as “plant trees for shade during extreme heat” and “encourage people to plant one tree for every household.” Under water and sanitation, participants called for better infrastructure and cooling features, including a “continuous flow of water through our taps” and public “fountains or ponds to cool the air.” The theme of a clean and healthy environment focused on waste management and hygiene, with recommendations like “provide bins on the streets to prevent pollution” and keep areas “free of stagnant water and bushy grass.” Health education and services emphasized awareness and local care, with calls to “create more awareness on health and environmental issues” and “build a well-equipped health center.” Lastly, civic engagement highlighted the role of young people in city upkeep, suggesting efforts to “mobilize the youth to clean the community” and “create sustainable jobs and education for the youth.”

## Discussion

### Summary of Findings

Our findings indicate that heatwaves are associated with a higher prevalence of self-reported health symptoms such as low mood, anxiety/stress, and headaches. For example, participants in Port Harcourt and Manila reported higher rates of symptoms during heatwaves, while Dar es Salaam showed less variation between heatwave and non-heatwave periods. Additionally, heatwaves appear to exacerbate disruptions in daily activities, particularly not enough food and the need for additional family support. These disruptions were most pronounced in Port Harcourt, where over half of participants reported not having enough food and relying more on family support during heatwaves. By contrast, other cities reported lower but still notable increases in these disruptions.

Beyond quantifiable impacts, participants expressed insights into how young people and caregivers imagine healthier, more resilient urban futures. Through thematic analysis of open-ended responses, five core priorities emerged: urban greening, improved sanitation, cleaner environments, better health education and services, and civic engagement. While these themes were common across all locations, some differences emerged. For instance, respondents in Accra and Kumasi emphasized sanitation and water, while those in Manila highlighted environmental cleanliness and waste management.

### Interpretations of results

The impacts of heatwaves are not evenly distributed. Younger participants and those with lower incomes reported more health symptoms and greater disruptions to daily life (Tables 1 & 2). This is consistent with research showing that children are physiologically more vulnerable to heatwaves due to their more limited ability to regulate body temperature (2,7). Younger people are also vulnerable to psychosocial factors. People with fewer resources are also less likely to have access to cooling, healthcare, or flexible work arrangements, which can protect against heat-related effects (3,14,15).

There were also differences between cities. For instance, compared to Accra, participants in Dar es Salaam and Ouagadougou consistently showed lower odds of reporting low mood and anxiety/stress. This may be due to a combination of intense heat, limited green space, crowded living conditions, and weak social support systems (16,17) in Accra. These results point to the need for local responses that address specific environmental and social conditions.

Across all pooled cities, participants reported more low mood, anxiety, stress, and headaches during heatwaves. These findings are in line with existing studies that link heat to mental health problems (18). Possible explanations include changes in brain chemistry and worsening of existing mental health conditions (18,19). Headaches may be a sign of physical stress or dehydration caused by heat (20).

Overall, well-being and sleep quality did not change significantly between heatwave and non-heatwave periods. This may reflect adaptation or coping strategies, such as adjusting daily routines or using fans and air conditioners where available (21,22) or cultural sleep practices. These findings differ from those of (23,24), who found that heatwaves worsened sleep. The increase in physical activity during heatwaves likely reflects work-related demands, particularly in urban settings where a significant proportion of the population is engaged in informal or outdoor labour, often without the option to reduce work hours or move indoors. This stands in contrast to findings from wealthier, service-based economies, where activity levels tend to decline during extreme heat due to greater access to indoor workspaces, flexible schedules, and climate-controlled environments (21,25,26).

The findings suggest that both not having enough food and the need for family support increased during heatwaves, particularly among younger, lower-income and participants in Port Harcourt. These outcomes are likely linked to the additional strain extreme heat places on daily life, reducing income-earning capacity, disrupting food storage and preparation, and increasing caregiving needs (2,14,27). Younger individuals may be more affected due to their greater reliance on caregivers, while low-income households face greater difficulty absorbing the financial and logistical impacts of heatwaves to access food and provide family support (2,6). These patterns align with existing studies showing that heatwaves can intensify economic and social vulnerabilities in cities, especially where public infrastructure and services are limited (2,6,14,27).

These community-derived themes closely align with global frameworks like the WHO Healthy Cities initiative and UNICEF’s climate resilience priorities for children (28,29), reinforcing the legitimacy and relevance of youth-informed planning in urban adaptation.

The perception of satisfaction with city responses should be interpreted with caution. In some contexts, this may reflect psychological resilience or adaptive preferences; in others, it could be a sign of normalized deprivation, where poor infrastructure is expected rather than contested (14). Perceived satisfaction with city interventions may be partly attributable to visible mitigation efforts, such as emergency alerts, cooling centers, and other adaptive infrastructure (21). In many cities, acceptance of city responses may reflect low expectations shaped by ongoing exposure and inadequate public services, resulting in household-level adaptations (14).

### Strengths and Limitations of the Study

This study has several strengths. It included participants from six diverse cities, which allowed us to compare experiences of CYP across different urban environments. The use of real-time temperature data and an automated system to identify heatwave periods helped ensure that we captured event periods consistently and accurately. The sample size was large and included a wide range of ages, income levels, which adds depth to the findings. By combining survey data with open-ended responses, the study not only measured the impact of heatwaves but also gave space for participants to share their ideas for healthier cities. The involvement of a Youth Advisory Group in designing the survey and shaping the analysis added further value and ensured the research remained grounded in the experiences of young people.

At the same time, there are some limitations. Online recruitment likely excluded individuals without consistent internet access or digital literacy, resulting in a sample that may overrepresent more connected, urban, or tech-savvy individuals. This selection bias may have led to an underrepresentation of those from lower-income or more vulnerable communities, potentially underestimating the true impact of heatwaves on these groups. While the survey was carefully translated and reviewed for clarity, cultural and linguistic differences in how questions were understood may have influenced participants’ responses. This may have introduced measurement bias, particularly for subjective responses such as mood, stress, or daily disruptions, and could affect the comparability of results across event periods. As a result, some observed differences may reflect variation in reporting styles rather than actual differences in experience.

Additionally, the cross-sectional design of the study limits the ability to infer causality. Because participants surveyed during heatwaves and non-heatwaves were not necessarily the same individuals, observed differences in health symptoms or daily disruptions may reflect variations in respondent characteristics rather than the effects of heatwave exposure itself. For example, differences in age, income, or location between the two samples may introduce confounding. In Manila, the sample during the heatwave was older on average than during the non-heatwave period. Such demographic shifts may help explain unexpected findings, such as lower reported rates of heat exhaustion during heatwaves, raising the possibility that some results reflect sample composition rather than true changes in experience.

Finally, sample sizes varied across cities and were particularly limited during the non-heatwave period, likely due to response fatigue among participants who had already completed the heatwave survey. These smaller and uneven samples may have influenced the findings, which are likely to reflect more strongly the experiences of cities with higher response rates.

### Implications for Policy and Practice

Our findings illustrate the need for climate-sensitive urban planning that centers the voices of CYP. Policy interventions should prioritize green infrastructure, equitable access to water and sanitation, and support systems for families during climate shocks. The expressed willingness of youth to engage in civic actions suggests untapped potential for co-creating resilient cities through participatory governance.

Urban heatwave preparedness must move beyond physical infrastructure to include social protection, mental health support, and climate education tailored to young urban populations. These findings align with existing global frameworks, including the WHO Healthy Cities initiative and UNICEF’s climate resilience priorities for children (28,29), reinforcing the legitimacy and relevance of youth-informed planning in urban climate adaptation. For policy-makers, this alignment strengthens the case for funding mechanisms that specifically support youth-led or community-driven adaptation efforts, ensuring that investments reflect the lived realities and priorities of those most affected by climate risks.

### Recommendations for Future Research

Future research should consider longitudinal designs that follow the same participants over time to better assess whether changes in reported health outcomes or daily disruptions are linked to heatwave exposure or differences in sample composition. Combining online recruitment with offline or community-based approaches would improve inclusion of individuals with limited internet access, who may be more vulnerable to extreme heat. Collecting data across multiple heatwave and non-heatwave periods could help increase sample sizes and enable more balanced comparisons across cities. Additionally, detailed city-specific analyses, along with an examination of within-city differences, such as demographics, or access to services, would provide a clearer understanding of how heatwaves affect health, wellbeing, and daily life. Finally, integrating tools such as wearable devices, mobile apps, or local temperature sensors could improve exposure measurement and enhance the accuracy of findings.

## Conclusion

This study demonstrates the significant and uneven impacts of heatwaves on CYP across diverse city contexts. Vulnerability was shaped not only by exposure to extreme heat, but also by intersecting factors such as age, socioeconomic status and location. While heatwaves were associated with heightened health symptoms, as well as daily disruptions, participants also expressed clear, community-based priorities for enhancing city resilience.

These findings highlight the need for youth-informed, context-specific adaptation strategies that recognize the lived experiences of young people in rapidly urbanizing settings. Investments in green infrastructure, accessible public health systems, and participatory governance can contribute to more equitable, healthy, and climate-resilient urban futures.

By centering the experiences of young people in fast-growing cities in the Global South, this study contributes to the growing evidence base on climate and health. It reinforces the link between heatwaves and adverse health outcomes, while also emphasizing the critical role of inclusive, city-specific solutions in addressing climate vulnerability.

## Data Availability

All anonymised datasets, data collection tools, and analysis code are publicly available via the London School of Hygiene & Tropical Medicine (LSHTM) Data Compass repository. The corresponding DataCite DOIs are included in the Supporting Information files. All other relevant materials are also provided as Supporting Information.

https://datacompass.lshtm.ac.uk/id/eprint/4298/

https://datacompass.lshtm.ac.uk/id/eprint/4617/

https://datacompass.lshtm.ac.uk/id/eprint/4615/

https://datacompass.lshtm.ac.uk/id/eprint/4616/

https://datacompass.lshtm.ac.uk/id/eprint/4653/

https://datacompass.lshtm.ac.uk/id/eprint/4689/

## Acknowledgments

We are sincerely grateful to the CYP and parents who took part in the real-time survey activities. We also extend our appreciation to the local teams and partner organizations across the participating cities for their role in facilitating data collection. Special thanks to the Empower Agency for their valuable contribution to the recruitment process and for providing training to staff involved in online data collection.

## Supporting information

**S1 File. Protocol for selecting the CCC Action Lab focal cities.** Describes the criteria and process used to identify and select the 178 focal cities used for the event identification in the study.

**S2 File. Advertisements for the six cities.** Includes screenshots and copies of the online recruitment advertisements used in each of the six study cities.

**S3 File. Data collection tools.** Presents the questionnaires in 12 languages used in the study.

**S4 File. Quantitative Dataset.** Contains the anonymized, cleaned dataset used for quantitative analysis in this study.

**S5 File. Data analysis plan.** Outlines the planned analytical approach for both quantitative and qualitative data.

**S6 File. Sample mayor’s email.** Provides an example of the outreach email used to inform city officials of the data collection in their city

**S1 Fig. Event sampling flowchart.** Illustrates the process for identifying of eligible heatwave and non-heatwave periods and participant recruitment in the six cities.

**S1 Table. Sample size calculation per city.** Displays the estimated minimum number of responses required per city for robust analysis.

**S2 Table. Advertisement performance and survey engagement.** Summarizes key metrics on ad impressions, click-throughs, and completed surveys by city.

**S3 Table. Demographic characteristics.** Provides a breakdown of age, gender, and parental status for survey respondents.

**S4 Table. Well-being (feeling today and sleep quality).** Presents participant-reported measures of general well-being and sleep quality.

**S5 Table. Health symptoms.** Lists the self-reported physical and emotional health symptoms during heatwave and non-heatwave periods.

**S6 Table. Logistic regression results for health symptoms during heatwaves.** Shows adjusted odds ratios for the likelihood of experiencing health symptoms during heatwave periods.

**S7 Table. Physical activity.** Summarizes reported levels and changes in physical activity during extreme heat periods.

**S8 Table. Disruptions in daily activities.** Details interruptions to school, work, and caregiving activities during heatwaves.

**S9 Table. Logistic regression results for daily activity disruptions during heatwaves.** Presents adjusted models assessing the association between heatwaves and daily activity disruptions.

**S10 Table. Concern about heatwaves.** Displays participants’ levels of concern about current and future heatwaves.

**S11 Table. Satisfaction with heatwave response.** Summarizes Children, Young People and Parents perspectives on their satisfaction on how well their cities responded to recent heatwaves.

**S12 Table. Thematic codebook and coded qualitative dataset.** Provides the codebook used for qualitative analysis and an excerpt of coded text illustrating key themes.

